# Inflammation and autoimmunity are interrelated in patients with sickle cell disease at a steady-state condition: implications for vaso-occlusive crisis, pain, and sensory sensitivity

**DOI:** 10.1101/2023.09.03.23294996

**Authors:** Wei Li, Andrew Q Pucka, Candice Debats, Brandon Reyes, Fahim Syed, Andrew R O’Brien, Rakesh Mehta, Naveen Manchanda, Seethal A Jacob, Brandon M Hardesty, Anne Greist, Steven E Harte, Richard E Harris, Qigui Yu, Ying Wang

## Abstract

This study aimed to comprehensively analyze inflammatory and autoimmune characteristics of patients with sickle cell disease (SCD) at a steady-state condition (StSt) compared to healthy controls (HCs) to explore the pathogenesis of StSt and its impact on patients’ well-being. The study cohort consisted of 40 StSt participants and 23 HCs enrolled between July 2021 and April 2023. StSt participants showed elevated white blood cell (WBC) counts and altered hematological measurements when compared to HCs. A multiplex immunoassay was used to profile 80 inflammatory cytokines/chemokines/growth factors in plasma samples from these SCD participants and HCs. Significantly higher plasma levels of 37 analytes were observed in SCD participants, with HGF, IL-18, IP-10, and MCP-2 being among the most significantly affected analytes. Additionally, autoantibody profiles were also altered, with elevated levels of anti-SSA/Ro60, anti-Ribosomal P, anti-Myeloperoxidase (MPO), and anti-PM/Scl-100 observed in SCD participants. Flow cytometric analysis revealed higher rates of red blood cell (RBC)/reticulocyte-leukocyte aggregation in SCD participants, predominantly involving monocytes. Notably, correlation analysis identified associations between inflammatory mediator levels, autoantibodies, RBC/reticulocyte-leukocyte aggregation, clinical lab test results, and pain crisis/sensitivity, shedding light on the intricate interactions between these factors. The findings underscore the potential significance of specific biomarkers and therapeutic targets that may hold promise for future investigations and clinical interventions tailored to the unique challenges posed by SCD. In addition, the correlations between vaso-occlusive crisis (VOC)/pain/sensory sensitivity and inflammation/immune dysregulation offer valuable insights into the pathogenesis of SCD and may lead to more targeted and effective therapeutic strategies.

## Introduction

Sickle cell disease (SCD) is a lifelong illness that affects multiple organ systems and can cause a range of complications, including acute and chronic pain, anemia, stroke, pulmonary hypertension, and organ damage (1, 2). The clinical manifestations of SCD can be broadly categorized into two phases: the steady-state (StSt) phase, characterized by mild to no symptoms associated with chronic hemolysis and persistent pain (3), and the severe pain and other clinical complications in the acute hemolytic/vaso-occlusive crisis (VOC) phase (4, 5). The acute VOC phase is marked by the sudden onset of severe pain, possibly accompanied with acute chest syndrome and stroke due to the blockage of small blood vessels by sickle-shaped red blood cells (RBCs). Individuals with SCD requiring high doses of opioids to manage VOC confront a series of substantial risks, such as an increased vulnerability to overdose and mortality, the potential development of opioid-induced hyperalgesia, and a compromised quality of life (QoL). Addressing these challenges becomes paramount, emphasizing the urgent need for evidence-based, effective, and safe pain management therapies tailored specifically for SCD. Indeed, pain is the hallmark of SCD, manifesting anywhere in the body and profoundly impacting patients’ QoL (6-8). SCD pain can manifest as acute recurrent painful crises associated with VOCs as well as chronic pain with or without nerve damage (7). The painful crisis, which evolves through four phases—prodromal, initial, established, and resolving—is a leading cause of hospitalization and emergency department treatments in SCD (7).

SCD is characterized by the presence of sickle-shaped RBCs with altered biophysical and biochemical properties (9). Unlike normal RBCs, which have a biconcave disc shape for flexibility and deformability, sickle-shaped RBCs lose their ability to deform and navigate through small blood vessels efficiently, leading to blockages and reduced blood flow. To compensate for the RBC loss, bone marrow produces more immature RBCs known as reticulocytes, releasing them into circulation and leading to elevated reticulocyte levels in the blood. In SCD participants, reticulocytes express higher levels of surface adhesion molecules such as Lutheran/basal cell adhesion molecule (Lu/BCAM) and alpha-4 beta-1 (α4β1) integrin (10-15). These adhesion molecules facilitate the binding of reticulocytes to the endothelium lining the blood vessels, causing activation of endothelial cells (ECs) and the release of inflammatory molecules (16-18). Moreover, reticulocytes in SCD interact with circulating leukocytes, including polymorphonuclear neutrophils (PMNs) and monocytes, leading to the formation of aggregates and increasing the occurrence of VOCs (19-21). These interactions are primarily mediated by α4β1 integrin and Lu/BCAM, reinforcing their adhesion to the endothelium (9, 22). *In vitro* studies have demonstrated that sickle-shaped RBCs can also bind to peripheral leukocytes, especially monocytes, via erythroid LW/ICAM-4 and CD44 receptors. These RBC/reticulocyte-leukocyte aggregates interact with ECs, resulting in EC activation (21). Thus, SCD is associated with increased adhesion of RBC/reticulocytes to leukocytes and ECs, which contribute to the complex pathophysiology underlying SCD pain (10-18).

Chronic inflammation persists in SCD due to continuous activation of immune cells or autoimmune responses triggered by various factors, such as hemolysis, vaso-occlusion, and sickle-shaped RBC aggregates. Hemolysis is a primary inflammatory trigger in SCD (17). Sickle-shaped RBCs are more fragile and prone to rupture, leading to the chronic release of hemoglobin and other cellular components. Hemoglobin is a potent pro-inflammatory molecule that activates immune cells such as monocytes, macrophages, and dendritic cells (DCs), prompting them to produce pro-inflammatory cytokines such as IL-1β (23), IL-6 (24), and TNF-α (23). Additionally, heme, an iron-containing component of hemoglobin, can further induce the production of pro-inflammatory cytokines, chemokines, and adhesion molecules by activating NF-*κ*B and TLR4 signaling pathways, and promote the recruitment of leukocytes and platelets to sites of inflammation, exacerbating the inflammatory response (25). In addition, vaso-occlusion of small blood vessels by sickle-shaped RBCs leads to tissue ischemia and subsequent hypoxia, activating ECs. The activated ECs produce and release pro-inflammatory cytokines and adhesion molecules (26, 27), which recruit lymphocytes to the site of inflammation, further perpetuating the inflammatory response. Furthermore, chronic activation of the coagulation system in SCD participants results in the release of pro-inflammatory mediators such as thrombin, which can activate ECs and promote the recruitment of inflammatory cells. SCD is also associated with oxidative stress, which contributes to inflammation by activating the NF-*κ*B pathway, leading to the production of pro-inflammatory cytokines and chemokines. Finally, the impact of autoantibodies on inflammation in SCD participants is noteworthy. Chronic hemolysis and cell damage release intracellular components like hemoglobin, heme, and cellular debris into the bloodstream, triggering the production of autoantibodies. These autoantibodies exacerbate inflammation by forming immune complexes that activate pro-inflammatory pathways, recruit immune cells, and contribute to tissue damage. Highlighting the complex pathophysiology that contributes to inflammation in SCD, a recent study quantified serum levels of 27 inflammatory cytokines, chemokine, and growth factors in 27 individuals in StSt, 22 individuals in VOC, and 53 healthy individuals (28). The study found that both pro- and anti-inflammatory cytokines are involved in the inflammatory response during SCD, regardless of clinical phase (28). This suggests that the dysregulation of cytokine production and balance may play a role in the disease’s pathophysiology.

The current study represents the initial phase of an ongoing clinical trial (ClinicalTrials.gov, NCT05045820) focused on investigating the clinical efficacy and neurobiological mechanisms of acupuncture analgesia in SCD participants, for which recruitment is ongoing. The primary objective of the current study was to characterize the pain and the underlying immune and inflammatory abnormalities in SCD participants. To achieve this objective, we examined plasma levels of inflammatory cytokines, chemokines, growth factors, soluble receptors, and effector molecules, as well as the profiles of autoantibodies and the aggregation between RBCs/reticulocytes and leukocytes. Moreover, we investigated the association between these analytes and VOCs, patient reported pain- and QoL-related outcomes, and sensory sensitivity. Our long-term goal is to identify potential mechanisms and therapeutic targets in SCD, gaining an improved understanding of the mechanisms behind acupuncture intervention.

## Materials and Methods

### Study participants

This work constitutes an initial part of an ongoing randomized clinical trial in SCD that commenced on June 29, 2021, and is scheduled to conclude on May 31, 2026. The primary inclusion criteria for participant enrollment included: 1) has been diagnosed with SCD, 2) experiencing chronic pain within the last 6 months or encountered at least one VOC within the past 12 months, 3) no recent changes in stimulant medication dosage or initiation, 4) willingness to continue their ongoing treatments, and 5) an agreement to limit using any new medications or treatment methods for pain management throughout the study. The major exclusion criteria included: 1) individuals with COVID-19 suspected or confirmed, 2) recent or ongoing pain management using acupuncture or acupuncture-related techniques within the last 6-months, and 3) presence of a concurrent autoimmune or inflammatory disease such as rheumatoid arthritis, systemic lupus erythematosus (SLE), and inflammatory bowel disease. In addition, participants who received a blood transfusion within the 90 days prior to recruitment were excluded for analyses. Age-, gender-, and ethnicity-matched health subjects without SCD were recruited as healthy controls (HCs). Detailed information regarding the inclusion and exclusion criteria can be found on ClinicalTrials.gov (NCT05045820).

Peripheral blood samples were collected from 40 SCD participants, comprising 17 males and 23 females, with ages ranging from 14 to 73 years. All participants were Black/African American. SCD participants and HCs were enrolled in this study through Indiana University Health hospitals in Indianapolis, the Indiana Hemophilia & Thrombosis Center, community hospitals, and other resources between July 2021 to April 2023. Peripheral blood was collected at StSt phase in heparin-coated tubes (BD Biosciences, Franklin Lakes, NJ) and subsequently separated into plasma and peripheral blood mononuclear cells (PBMCs). PBMCs were either directly used or cryopreserved in liquid nitrogen until use. Plasma samples were stored at −80°C until use. To establish a comparison, we included PBMC and plasma samples from 23 healthy volunteers, matched in terms of age, sex, and race, to serve as HCs. Detailed demographic and clinical characteristics of both SCD participants and HCs are summarized in Table 1. This study was performed with the approval of the Institutional Review Boards (IRB) at Indiana University School of Medicine, and each participant provided written informed consent during the screening visit prior to the subsequent study procedure.

**Table 1.** Clinical and hematological characteristics of the study participants.

| Parameters | HCs (n=23) | SCD (n=40) | p value |
| --- | --- | --- | --- |
| **Demographics** |  |  |  |
| Age (years) | 38 (22-57) | 34 (23-41) | 0.16 |
| Gender (% females) | 13 (57%) | 23 (58%) | 0.94 |
| SS/Sβ0/SC/Sβ+ thalassemia (n/n/n/n) | N/A | 23/3/11/3 |  |
| **Hemogram/Platelets/WBC Differential** |  |  |  |
| WBC (k/cumm) | 5.1 (4.0-6.3) | 8.8 (6.1-11.4) | **0.0001** |
| RBC (million/cumm) | 4.6 (4.3-5.1) | 2.9 (2.4-3.7) | **<0.0001** |
| Hgb (GM/dL) | 13.0 (12.2-14.3) | 10.2 (8.0-11.6) | **<0.0001** |
| HCT (%) | 38.8 (36.4-41.9) | 29.2 (23.6-34.0) | **<0.0001** |
| Absolute Retic Number (k/cumm) | 48.8 (41.6-71.0) | 131.2 (88.5-216.4) | **<0.0001** |
| Hemoglobin A (%) | 97.4 (88.5-97.6) | 23.0 (13.8-30.6) | **<0.0001** |
| Hemoglobin S (%) |  | 63.4 (49.1-73.1) |  |
| Hemoglobin F (%) |  | 5.8 (2.4-13.4) |  |
| MCV (fL) [81-99] | 86 (81-89) | 94 (83-104) | **0.020** |
| MCH (pg) [27.0-34.0] | 28.4 (27.3-29.6) | 32.3 (27.5-35.8) | **0.0033** |
| RDW (%) [11.5-14.5] | 13.7 (13.3-14.9) | 18.4 (15.8-23.6) | **<0.0001** |
| Platelet (k/cumm) [150-450] | 246 (231-270) | 309 (247-424) | **0.0029** |
| Absolute Neutrophil | 2.75 (1.6-3.8) | 4.8 (3.1-7.2) | **0.0005** |
| Absolute Lymphocyte | 2.1 (1.5-2.5) | 2.4 (1.7-4.1) | 0.055<sup>#</sup> |
| Absolute Monocyte | 0.4 (0.3-0.4) | 0.8 (0.5-1.1) | **<0.0001** |
| Absolute Eosinophil | 0.1 (0.1-0.2) | 0.2 (0.1-0.4) | 0.058<sup>#</sup> |
| Absolute Basophil | 0 (0-0) | 0.1 (0-0.1) | **0.0007** |
| **Immunological tests** |  |  |  |
| C-reactive protein (mg/ml) | 1.1 (0.4-2.7) | 3.9 (1.6-7.3) | **0.0038** |

**Note:** HCs, healthy controls; SCD, sickle cell disease; WBC, white blood cell; RBC, red blood cell; Hgb, hemoglobin; HCT, hematocrit; MCV, mean corpuscular volume; MCH, mean corpuscular hemoglobin; RDW, red cell distribution width. The Mann-Whitney test was used to compare differences between patients with SCD and healthy controls (HC) for continuous variables. $\chi^2$ test was used for comparison of gender distribution between the 2 groups. $p < 0.05$ was considered significant. <sup>#</sup> $p < 0.06$ was considered as a clear trend toward significance.

### Patient-reported outcome measures (PROMs)

The Patient-Reported Outcomes Measurement Information System (PROMIS)-29 Questionnaire was used to evaluate pain intensity and interference, as well as physical function (29). Neuropathic pain symptoms were evaluated using the PainDETECT Questionnaire (higher score indicated higher pain) (30, 31). The Widespread Pain Index was used to evaluate the spatial distribution of pain across the body (higher score indicated higher pain) (32, 33). In addition, depression was evaluated using the Hospital Anxiety and Depression Scale (HADS) (higher score indicated higher depression) (34). Physical function was assessed with PROMIS-29 (higher score indicated more physical dysfunction) (29). Pain-related QoL was evaluated using the Pediatric Quality of Life Inventory (PedsQL) targeting 3 different age groups (13-18, 18-25, or 25+ years old), with higher scores representing better QOL (35). The number of patient-reported VOCs in the preceding 12 months was documented (36, 37). The number of days between blood draw and the most recent or future VOCs was recorded as the time intervals after or before VOCs.

### Quantitative Sensory Testing (QST)

QST is a well-established experimental protocol designed to investigate both ascending excitatory and descending inhibitory aspects of pain processing by assessing an individual’s perceptual response to various stimuli (38). QST was performed at up to three different body sites, including the primary testing site(s), which was the area(s) reported as most painful by each patient, along with the dominant-side ventral forearm and/or the dominant-side upper trapezius muscle, as described in previous studies on SCD (38, 39). HCs received primary testing at sites matched with those identified in patients, in addition to testing at the dominant forearm and/or trapezius.

Thermal (heat/cold) Detection/Pain Threshold was determined at each testing site using a TCA11 (QST-Lab, Strasbourg, France) with a thermal probe in contact with the subject’s skin surface. The thermode temperature was gradually adjusted from a baseline temperature at a controlled rate of 0.5 - 1 °C/s. Subjects indicated the thermal detection threshold (when they first felt the thermal stimulus) and the hot pain threshold (when they first felt pain from the thermal stimulus). The average of three trials for each test was used for analysis.

Mechanical Detection Threshold (MDT)/Mechanical Pain Threshold (MPT) were examined using von Frey monofilaments (Stoelting, Wood Dale, IL) and calibrated pinprick stimuli (MRC Systems GmbH, Heidelberg, Germany) respectively. Each von Frey monofilament was applied three times in ascending sequence until the stimulus was detected in at least two out of three trials. The next lower von Frey monofilament was then applied, and the lowest filament to be detected at least twice was considered the detection threshold. The MPT was determined using different pinprick probes applied to the skin surface of each site. Testing started with a stimulation intensity of 8 mN and in each case, the next higher pinprick stimulator was applied until the perception of “touch” changed its quality towards an additional percept of “sharp”, “pricking,” or “stinging.” The corresponding intensity represented the first suprathreshold value. Once the first painful stimulus was perceived, the testing direction was changed step-wise towards lower stimulus intensities until the first stimulus perceived as “blunt” and no longer as being “sharp,” “pricking” or “stinging” (subthreshold value). Again, a directional change towards higher intensities occurred and the cycle was repeated until five suprathreshold and five subthreshold values were determined. An inflection point was calculated as the average value of the ten suprathreshold and subthreshold to determine the MPT.

Mechanical Temporal Summation (MTS) was assessed using a single 256 mN pinprick (MRC Systems GmbH, Heidelberg, Germany) stimulus applied in triplicate to the skin surface of the selected sites, followed by a series of 10 identical stimuli (1 Hz – metronome-guided). MTS was calculated as the average pain rating from the series of 10 stimuli minus the average pain rating from the three trials with the single stimulus.

Pressure Pain Threshold (PPT)/Pressure Pain Tolerance (PPTol) was assessed using a digital, handheld pressure algometer (Algometer II, Somedic SenseLab AB, Norra Mellby, Sweden). The pressure was manually increased at a rate of 50 kPa/s (1000 kPa max) until participants indicated that the sensation of pressure became one of faint pain (PPT) and the maximum pressure pain that the participant can tolerate (PPTol), respectively. The average of 3 trials per site was used for analysis.

Conditioned Pain Modulation (CPM): Tonic pressure pain was used as the conditioning stimulus delivered via a cuff (Hokanson, Bellevue, WA) attached to the gastrocnemius muscle of the non-dominant leg. Pressure intensities were individually calibrated for each participant to elicit moderate pain (pain rating at 40-60 on a scale of 100) (40, 41). PPT served as the test stimulus and was measured 3 times at the dominant trapezius muscle prior to and during the cuff stimulation. Pain ratings were obtained every 15 s prior to and during cuff stimulation for up to 90s. CPM magnitude was calculated as patient-reported pain ratings during conditioned cuff pressure stimuli.

Only QST results on the standard sites (forearm, trapezius) were used for correlation analysis as each SCD participant often had different primary painful site(s).

### Multiplex immunoassays and enzyme-linked immunosorbent assay (ELISA)

Plasma concentrations of 80 inflammatory human cytokines, chemokines, growth factors, soluble receptors, and effector molecules and 18 human autoantibodies were simultaneously measured using the Immune Response 80-Plex Human ProcartaPlex™ Panel (Cat. #: EPX800-10080-901, Invitrogen, Carlsbad, CA) and the MILLIPLEX MAP Human Autoimmune Autoantibody Panel (HAIAB-10K, MilliporeSigma, Burlington, MA), respectively, according to the manufacturer’s instructions. The beads were read on a BioPlex 200 system (Bio-Rad, Hercules, CA). The standards at 4-fold serial dilutions were run on each plate in duplicate and used to calculate the concentrations of human cytokines, chemokines, growth factors, soluble receptors, and effector molecules using the Bio-Plex Manager Software (Bio-Rad, Hercules, CA) as previously reported (42). Plasma samples were diluted 100-fold for the autoantibody multiplex assay, and the levels of the autoantibodies were reported as mean fluorescence intensity (MFI) after background MFI subtraction. The plasma levels of C-reactive protein (CRP) were quantified using human CRP Duoset ELISA Kit (R&D Systems, Minneapolis, MN) according to the manufacturer’s instructions.

### Flow cytometry

Freshly prepared PBMCs were stained with fluorochrome-conjugated antibodies against human CD45, CD71 (expressed on erythroid precursors), BCAM, and CD235ab (also known as GPA: glycophorin A, the major sialoglycoprotein on RBCs and their precursors) to measure the aggregation between RBC/erythroid precursors and CD45^+^ leukocytes. Cells stained with surface markers were acquired using a BD LSRFortessa flow cytometer (BD Biosciences, San Jose, CA). Flow data were analyzed using FlowJo v10 software (Tree Star, San Carlos, CA). The percentage of CD235ab^hi^ cells among the CD45^+^ leukocytes was defined as the RBC/reticulocyte-leukocyte aggregation rate. The association of RBCs/reticulocytes with lymphocytes or monocytes were determined based on their characteristics of FSC and SSC on flow cytometry, respectively.

### Statistical analysis

Statistical analysis was performed using GraphPad Prism 10 and SPSS 29. Data were expressed as median and interquartile range (Tables 1-3). Differences between 2 groups were calculated using the Mann-Whitney test for continuous variables (Tables 1-2). χ 2 test was used for comparison between 2 groups for gender distribution (Table 1). Adjusted *p* values were calculated for the 80-plex and 18-plex analytes using the Holm-Šídák correction for multiple comparisons (Table 3). Inflammatory mediators that were heightened in the SCD participants were used for subsequent Spearman correlation analyses using age, gender and SCD genotype as covariables (Tables 4-6). *p* < 0.05 was considered statistically significant. ^#^*p* < 0.06 was considered a clear trend toward significance.

**Table 2.** Patient-reported outcome measures and quantitative sensory testing.

| Parameters | HCS (n=23) | SCD (n=33) | p value |
| --- | --- | --- | --- |
| **PROMs** |  |  |  |
| PainDetect_Total score | 7 (7-7) | 19 (14-24) | **<0.0001** |
| BPI_Pain Interference Score | 0 (0-0) | 4.2 (1.8-5.6) | **<0.0001** |
| FPS_Widespread Pain Index | 0 (0-0) | 6 (3-8) | **<0.0001** |
| PROMISE 29_Physical Function Score | 4 (4-4) | 7 (5-11.5) | **<0.0001** |
| PROMISE 29_Pain Intensity Score | 0 (0-0) | 5 (4-6) | **<0.0001** |
| HADS_Depression Score | 1 (0-3) | 4.5 (2.0-7.8) | **0.0002** |
| Pain Episode Frequency Score | N/A | 8 (6-9) |  |
| Number of VOCs in preceding 12 months | N/A | 4 (2-7) |  |
| PedsQL_Total Score | N/A | 52 (43-63) |  |
| **Quantitative Sensory Testing** |  |  |  |
| MDT (forearm) | 0.4 (0.2-0.5) | 0.3 (0.1-0.5) | **0.0433** |
| MDT (primary site) | 0.3 (0.2-0.5) | 0.2 (0.1-0.6) | 0.4946 |
| MPT (forearm) | 77.0 (40.0-105.7) | 62.4 (23.2-128.0) | 0.6220 |
| MPT (primary site) | 83 (45.8-161.7) | 41.6 (15.3-104.0) | **0.0444** |
| MTS (forearm) | 6.7 (0.0-29.2) | 8.7 (3.3-23.3) | 0.3783 |
| MTS (primary site) | 5.0 (1.5-18.8) | 15.3 (5.3-23.0) | **0.0434** |
| CDT (forearm) | -2.4 ((-4.3)-(-1.6)) | -2.0 ((-3.1)-(-1.5)) | 0.3888 |
| CDT (primary site) | -2.0 ((-3.4)-(-1.4)) | -2.4 ((-3.5)-(-1.5)) | 0.5940 |
| CPT (forearm) | -13.3 ((-24.6)-(-5.4)) | -6.8 ((-13.1)-(-3.9)) | **0.0276** |
| CPT (primary site) | -15.65 ((-26.5)-(-6)) | -7.3 ((-12.7)-(-4.3)) | **0.0274** |
| HDT (forearm) | 3.3 (2.5-4.0) | 3.6 (2.5-4.4) | 0.8099 |
| HDT (primary site) | 3.6 (3.1-5.9) | 4.1 (2.7-5.7) | 0.6730 |
| HPT (forearm) | 7.8 (4.8-11.1) | 8.7 (5.4-10.7) | 0.9278 |
| HPT (primary site) | 7.3 (6.2-10.9) | 8.3 (6.1-10.3) | 0.8623 |
| PPT (trapezius) | 255.5 (179.3-364.5) | 203.5 (164.0-278.5) | **0.0404** |
| PPT (primary site) | 321.0 (196.0-539.0) | 192.0 (142.0-263.8) | **0.0026** |
| PPTol (trapezius) | 415.5 (341.8-559.6) | 316.5 (225.0-391.8) | **0.0129** |
| PPTol (primary site) | 526.0 (316.0-827.0) | 302.0 (223.8-463.8) | **0.0029** |
| CPM (pain rating) | 43.0 (30.0-54.0) | 39.0 (28.0-51.0) | 0.7441 |

**Note:** HCs, healthy controls; SCD, sickle cell disease; MDT, mechanical detection threshold; MPT, mechanical pain threshold; MTS, mechanical temporal summation; Mechanical CDT, cold detection threshold; CPT, cold pain threshold; HDT, heat detection threshold; HPT, heat pain threshold; PPT, pressure pain threshold; PPTol, pressure pain tolerance; CPM, conditioned pain modulation. The Mann-Whitney test was used to compare differences between patients with SCD and HCs. $p < 0.05$ was considered significant.

**Table 3:** Upregulated inflammatory mediators in SCD subjects as detected by 80plex immunoassay.

| Categories | Analytes | HCs (n=18) | SCD (n=36) | *p* | Adjusted *p* |
| --- | --- | --- | --- | --- | --- |
| **Chemokines** | CCL21 | 0.1 (0-12.8) | 20.4 (0.6-71.5) | 0.00280 | 0.17337 |
|  | **CCL23** | 487 (302-639) | 874 (574-1285) | 0.00014 | **0.01046** |
|  | Eotaxin-2 | 0.1 (0-6.6) | 10.4 (0-97.7) | 0.04479 | 0.88393 |
|  | **Gal-3** | 6749 (4949-8982) | 15144 (6910-34355) | 0.00055 | **0.03958** |
| | GRO-α | 1.7 (1.3-2.0) | 2.4 (1.7-3.1) | 0.00625 | 0.33050 |
|  | **IP-10** | 17.2 (10.1-25.6) | 36.9 (21.3-60.7) | 0.00012 | **0.00891** |
|  | **MCP-2** | 0.2 (0.1-0.7) | 2.1 (0.7-5.3) | 0.00001 | **0.00091** |
| | MIP-1α | 0 (0-1.6) | 2.7 (0.7-6.9) | 0.00629 | 0.33050 |
| | MIP-1β | 30.0 (14.9-40.5) | 56.8 (29.7-71.5) | 0.00134 | 0.09067 |
| | MIP-3α | 5.1 (4.4-5.5) | 5.9 (5.5-7.6) | 0.00196 | 0.12821 |
| **Effectors** | Granzyme A | 3.4 (0-10.9) | 11.1 (4.6-23.5) | 0.00442 | 0.25324 |
|  | Granzyme B | 10.4 (7.0-28.0) | 20.2 (12.6-31.9) | 0.01758 | 0.62295 |
| **Growth factors** | BAFF | 3.053 (1.051-4.074) | 4.283 (2.555-6.08) | 0.01758 | 0.62295 |
|  | bNGF | 1.1 (0.9-3.1) | 2.1 (1.2-4.6) | 0.02762 | 0.75348 |
|  | G-CSF | 4.2 (0-89.4) | 50.7 (25.3-116.3) | 0.01299 | 0.51903 |
|  | GM-CSF | 0 (0-0) | 3.8 (0-20.5) | 0.00256 | 0.16236 |
|  | **HGF** | 6.4 (2.9-20.3) | 56.2 (13.1-70.2) | 0.00003 | **0.00253** |
|  | IL-7 | 0 (0-0) | 0 (0-1.1) | 0.02305 | 0.70251 |
|  | IL-20 | 6.9 (5.5-10.5) | 9.6 (6.7-21.8) | 0.02509 | 0.72631 |
|  | **IL-34** | 11.5 (8.3-18.5) | 21.9 (13.7-43.9) | 0.00051 | **0.03716** |
|  | LIF | 0.6 (0.4-2.7) | 2.4 (1.1-4.8) | 0.00991 | 0.44448 |
|  | VEGF-A | 50.02 (33.59-65.04) | 73.17 (35.73-156.1) | 0.0523<sup>#</sup> | 0.91076 |
| **Inflammatory cytokines** | **IFN-γ** | 1.8 (1.4-2.2) | 3.9 (2.3-4.8) | 0.00023 | **0.01737** |
| | IL-1α | 3.9 (2.5-11.0) | 6.9 (4.1-16.4) | 0.04832 | 0.89752 |
|  | IL-2 | 4.3 (3.2-6.2) | 9.1 (4.5-14.7) | 0.00940 | 0.43242 |
|  | IL-8 | 0.7 (0.1-6.9) | 5.5 (2.1-10.2) | 0.01183 | 0.49255 |
|  | IL-9 | 0.2 (0-1.4) | 1.4 (0.5-3.0) | 0.01134 | 0.48383 |
|  | IL-17A | 0 (0-1.725) | 0.98 (0-3.959) | 0.05482<sup>#</sup> | 0.91633 |
|  | **IL-18** | 5.0 (3.4-7.1) | 13.7 (7.2-50.8) | 0.00002 | **0.00173** |
| | TNF-α | 2.2 (1.9-2.6) | 2.7 (2.3-3.2) | 0.00460 | 0.25884 |
| | TNF-β | 0.1 (0-2.2) | 1.5 (0.4-4.8) | 0.02021 | 0.66116 |
| **Type II inflammatory cytokines** | IL-4 | 19.4 (11.2-35.1) | 37.3 (18.5-77.2) | 0.00901 | 0.42436 |
|  | IL-5 | 0 (0-0.2) | 3.5 (0-12.3) | 0.00290 | 0.17675 |
|  | IL-6 | 0 (0-0.8) | 1.5 (0-7.4) | 0.03843 | 0.84757 |
|  | TSLP | 1.3 (0.8-2.2) | 2.3 (1.7-3.4) | 0.00791 | 0.38881 |
| **Soluble receptors** | **PTX3** | 1137 (798-1924) | 3045 (1557-5283) | 0.00064 | **0.04533** |
|  | TREM-1 | 0 (0-235.4) | 248.2 (16.3-761.1) | 0.02832 | 0.75524 |

**Note:** HCs, healthy controls; SCD, sickle cell disease. The Mann-Whitney test was used to compare differences between patients with SCD and healthy controls (HC) without and with Holm-Šídák correction for multiple comparisons. $p < 0.05$ was considered significant. <sup>#</sup> $p < 0.06$ was considered a clear trend toward significance.

**Table 4.** Correlations of upregulated inflammatory mediators with altered autoantibodies and RBC/reticulocyte-leukocyte aggregation.

| Categories | Analytes | Autoantibodies |  | Aggregation |
| --- | --- | --- | --- | --- |
|  |  | SSA/Ro60 | Myeloperoxidase | % |
| Chemokines | CCL23<sup>&</sup> |  |  | 0.88* |
| | MIP-1α | | | 0.88* |
| | MIP-1β | | 0.37* | |
| Effector | Granzyme A | 0.34<sup>#</sup> |  |  |
| Growth factors | HGF<sup>&</sup> |  |  | 0.89* |
|  | IL-34<sup>&</sup> | 0.48** |  |  |
|  | VEGF-A | 0.33<sup>#</sup> |  |  |
| Proinflammatory cytokines | IL-1α | 0.35* | 0.39* | |
|  | IL-9 | 0.36* |  |  |
| Type II cytokines | IL-4 | 0.48** |  |  |
|  | IL-5 | 0.33<sup>#</sup> |  |  |

**Note:** Aggregation, RBC/reticulocyte-leukocyte aggregation. Spearman correlation analyses were performed including age, gender and SCD genotype as covariables. \* $p < 0.05$ ; \*\* $p < 0.01$ , \*\*\* $p < 0.001$ , # $p < 0.06$ , a clear trend toward significance, &mediators with adjusted $p < 0.05$ in Table 3.

## Results

### Characteristics of the study cohort: demographics, laboratory exams, patient reported outcomes and sensory sensitivity

Demographics and clinical characteristics of participants are summarized in Table 1. SCD participants and HCs showed no significant differences in terms of age and gender distribution. Compared to the HCs, SCD participants exhibited higher numbers of white blood cells (WBCs), reticulocytes, platelets, neutrophils, monocytes, and basophils but reduced RBCs, hemoglobin, and hematocrit (HCT). Additionally, SCD participants displayed elevated levels of mean corpuscular volume (MCV), mean corpuscular hemoglobin (MCH), red cell distribution width (RDW), and the systemic inflammation marker CRP than HCs (Table 1). The differences in the absolute numbers of lymphocytes and eosinophils approached a clear trend toward significance (p < 0.06) between the two groups (Table 1).

As indicated in Table 2, SCD participants reported increased pain interference and intensity, more painful body sites, reduced physical functioning, and higher levels of depression relative to HCs. Compared to HCs, SCD participants also displayed hypersensitivity to experimental mechanical (MDT, MTP, MTS), cold (CPT), and pressure stimuli (PPT and PPTol), but not hot stimuli (HDT, HPT) as measured by QST (Table 2).

### SCD participants presented with elevated proinflammatory cytokines, chemokines, growth factors, effector molecules, and soluble receptors compared to HCs

SCD participants had significantly higher plasma levels of 35 analytes compared to HCs (Table 3). These elevated analytes encompassed 10 chemokines (CCL21, CCL23, Eotaxin-2, Gal-3, GRO-α, IP-10, MCP-2, MIP-1α, MIP-1β, and MIP-3α), 2 effector molecules (granzymes A and B), 9 growth factors (BAFF, bNGF, G-CSF, GM-CSF, HGF, IL-7, IL-20, IL-34, and LIF), 8 pro-inflammatory cytokines (IFN-γ, IL-1α, IL-2, IL-8, IL-9, IL-18, TNF-α, and TNF-β), 4 type II inflammatory/anti-inflammatory cytokines (IL-4, IL-5, IL-6, and TSLP), and 2 immune modulating soluble receptors (PTX3 and TREM-1) (Table 3 and Supplementary Table 1). In addition, levels of the endothelial cell growth factor VEGF-A and the proinflammatory cytokine IL-17A trended higher in the SCD participants (*p* < 0.06) (Table 3 and Supplementary Table 1). After being corrected for multiple comparisons, 9 out of the 37 altered analytes remained significantly higher in the SCD participants compared to the HCs. These included 4 chemokines (CCL23, Gal-3, IP-10, and MCP-2), 2 growth factors (HGF and IL-34), 2 proinflammatory cytokines (IFN-g and IL-18), and the soluble receptor PTX3 (Table 3 and Supplementary Table 1). Notably, among these altered analytes, HGF, IL-18, IP-10, and MCP-2 exhibited the most significant upregulation in SCD participants in comparison to HCs (Table 3 and Supplementary Table 2).

### Both anti-nuclear autoantibodies (ANAs) and non-ANAs were present and elevated in SCD participants

Plasma concentrations of autoantibodies against 18 human antigens were simultaneously measured using a multiplex immunoassay. We identified the presence of autoantibodies against 4 human antigens, including Sjögren’s Syndrome-related antigen A/Ro60 kDa (SSA/Ro60), ribosomal P, myeloperoxidase (MPO), and PM/Scl-100, which were significantly elevated in SCD participants when compared to HCs (Fig. 1A, 1B). After multiple comparison corrections, SSA/Ro60 and MPO remained significantly higher in the SCD subjects (*p* = 0.048 and *p* = 0.022, respectively). It is noteworthy that while SSA/Ro60 autoantibody is typically classified as an ANA, autoantibodies against the ribosomal P, MPO, or PM/Scl-100 are not categorized as ANAs (non-ANAs). Thus, both ANAs and non-ANAs in the bloodstream were significantly elevated in SCD participants compared to HCs.

**Fig. 1.**
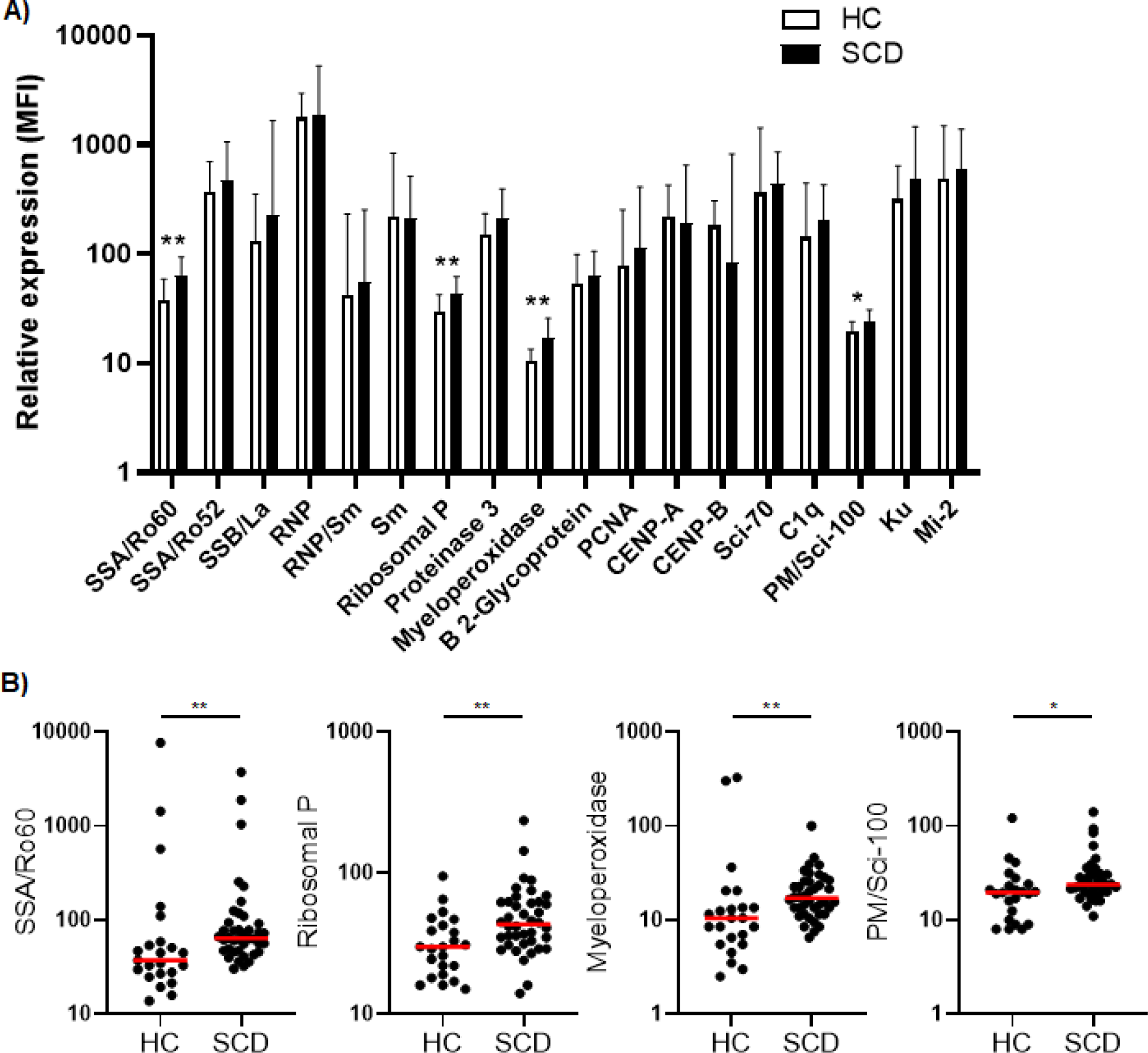
Comparative analysis of quantity and individual incidence of elevated autoantibodies in SCD participants versus healthy controls. **A)** The levels of plasma autoantibodies against 18 autoantigens in SCD participants (n=40) versus healthy controls (HCs, n=23). The median fluorescence intensity (MFI) after background MFI subtraction was used to represent relative expression of the 18 autoantibodies in SCD and HC samples, shown as median + upper interquartile range (IQR). **B)** Scatter plots demonstrating the plasma levels of 4 autoantibodies (anti-SSA/Ro60, anti-Ribosomal P, anti-Myeloperoxidase, and anti-PM/Scl-100) that were significantly elevated in SCD participants compared to healthy controls. Lines represent the median. SCD, sickle cell disease; HC, healthy control. Mann-Whitney test was used to compare the results from SCD patient versus HC samples. *\*p* < 0.05; *\*\*p* < 0.01.

### SCD participants exhibited abnormally high rates of RBC/reticulocyte-leukocyte aggregation

RBC/reticulocyte-leukocyte aggregates in SCD participants were assessed using flow cytometry (Fig. 2A). Freshly isolated PBMCs were stained with antibodies again human CD45 (a marker expressed on all leucocytes), CD235ab (a marker expressed on RBCs and their precursors), BCAM, and CD71 (transferring receptor-1, a specific marker of reticulocytes among the RBC population). As depicted in Fig. 2B, the percentage of total CD235ab^hi^CD45^+^ aggregates were significantly higher in SCD participants compared to HCs. The percentage of CD71^+^ reticulocyte aggregates was significantly higher in SCD participants compared to HCs (Fig. 2C, left panel). In parallel, the percentage of CD71^+^ reticulocytes in free fraction were also significantly higher in SCD participants compared to HCs, indicating an increase of reticulocytes in SCD participants (Fig. 2C, right panel). Similarly, BCAM expression on aggregates or unaggregated RBCs was significantly higher in SCD participants compared to HCs (Fig. 2D). This observation suggested that StSt PBMC samples contained higher numbers of RBCs or RBC precursors, such as reticulocytes, and higher levels of the cell adhesion molecule BCAM, thus contributing to the increased occurrence of aggregates. Compared to HCs, the percentage of CD45^+^ lymphocytes in the aggregates were significantly lower in SCD participants, whereas the frequency of monocytes was higher (Fig. 2E). When the immune cell types in PBMCs and the aggregates in the SCD participants were analyzed and compared, monocytes were enriched within the aggregates with a corresponding decrease of lymphocytes within aggregation relative to PBMCs (Fig. 2F), indicating RBCs/RBC precursors preferentially interacted with monocytes within the aggregates.

**Fig. 2.**
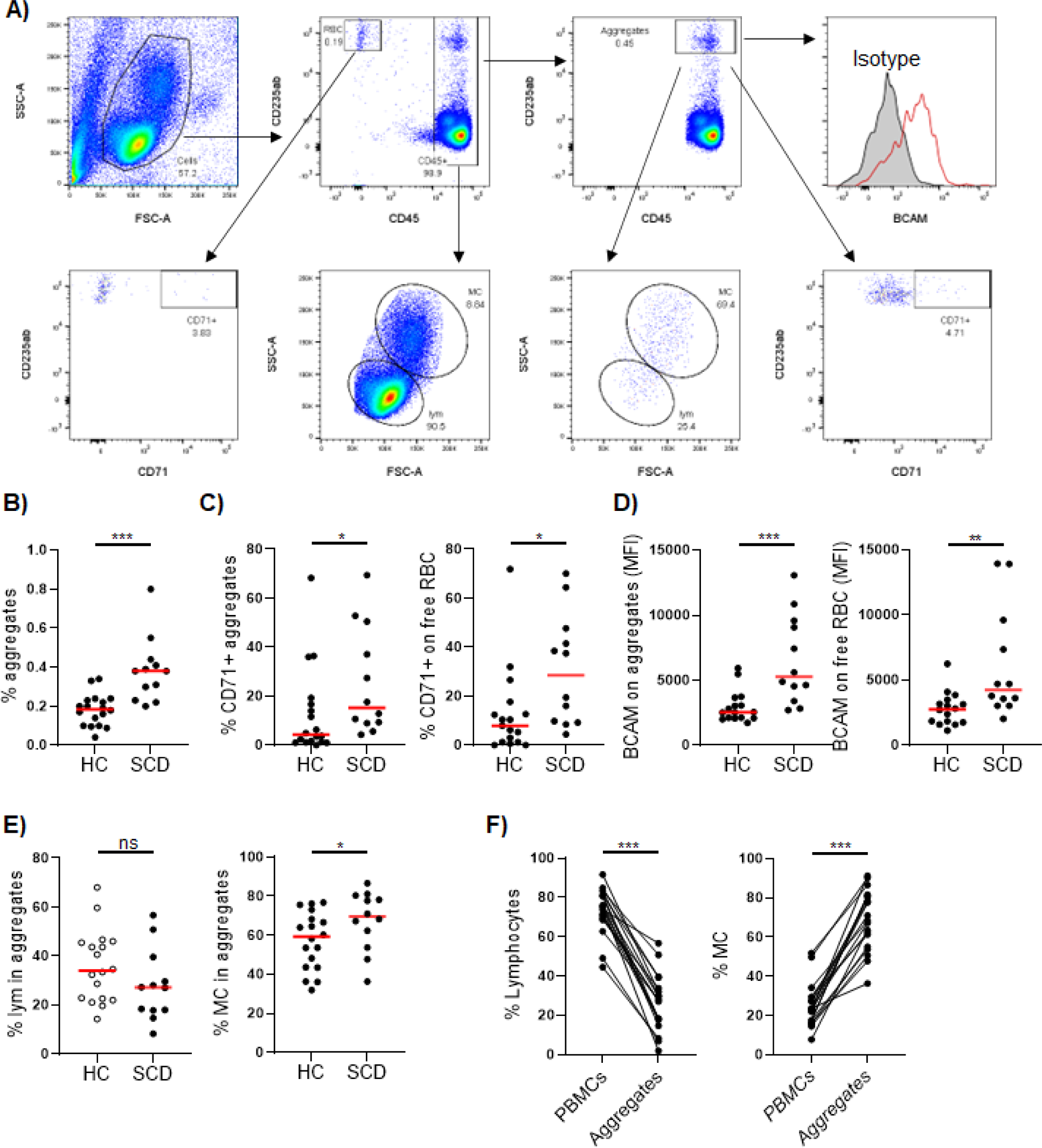
Comparison of RBC/reticulocyte aggregation with CD45^+^ PBMCs in SCD participants versus healthy controls. Freshly isolated peripheral blood mononuclear cells (PBMCs) from the SCD participants (n=12) and healthy controls (HC, n=17) were analyzed for RBC/reticulocyte aggregation with CD45^+^ PBMCs by flow cytometry. **A)** Gating strategies for flow cytometric analysis of RBC/reticulocyte-CD45^+^ leukocytes aggregation. PBMCs were stained with fluorochrome-conjugated antibodies against human CD45, CD235ab, CD71, BCAM and isotype control antibodies. CD235ab^+^ cells that were not associated with CD45^+^ PBMCs were defined as RBC/reticulocyte in free fraction (RBC). Within the CD45^+^ cells, CD235ab^hi^ cells were defined as the RBC/reticulocyte-CD45^+^ leukocyte aggregates. The gate for CD71^+^ RBC/reticulocyte and RBC/reticulocyte-leukocyte aggregates were based on the isotype control. BCAM level was expressed as mean fluorescent intensity (MFI). Lymphocytes (LYM) and monocytes (MC) were gated based on their FCS and SSC chrematistics. **B)** Scatter plots showing a significantly higher percentage of RBC/reticulocyte-CD45^+^ leukocyte aggregation in the SCD participants than HC. **C, D)** Scatter plots displaying higher levels of CD71^+^ erythroid precursors (C) and the cell adhesion molecule BCAM (D) on both aggregated and unaggregated RBC/reticulocyte in the SCD participants compared to HC. **E)** Scatter plots comparing the frequencies of lymphocytes and monocytes within RBC/reticulocyte-CD45^+^ leukocyte aggregates**. F)** Before and after plots showing the percentages of lymphocytes (left) were reduced while those of monocytes (right) were heightened in the aggregates compared to PBMCs in the SCD subjects. Mann-Whitney test was used to compare the results from SCD patient versus HC samples. Wilcoxon test was used to calculate the differences between aggregates and PBMCs. *\*p* < 0.05; *\*\*p* < 0.01; *\*\*\*p* < 0.001.

### Comprehensive analysis revealed correlations of inflammatory mediators with autoantibodies and RBC/reticulocyte-leukocyte aggregation in SCD participants

Next, we evaluated correlations between the 37 elevated inflammatory mediators (9 analytes with adjusted p<0.05 bolded) that were listed in Table 3, altered autoantibodies targeting SSA/Ro60, ribosomal PMPO, and PM/Scl-100, and RBC/reticulocyte-leukocyte aggregation in StSt participants (Table 4). The elevated plasma level of IL-4 and IL-5, both type II cytokines known for facilitating the development of B cells into antibody-producing plasma cells, exhibited significant positive associations with SSA/Ro60 autoantibodies. These autoantibodies are a type of ANAs commonly linked to autoimmune diseases such as SLE (Table 4) (43). IL-4 also exhibited a trend towards correlation with ribosomal P and MPO autoantibodies (*r* = 0.31, *p* = 0.076 for both). Additionally, SSA/Ro60 autoantibodies showed associations with multiple inflammatory mediators, including the effector molecule granzyme A, 2 growth factors (IL-34 and VEGF-A), and 2 proinflammatory cytokines (IL-1a and IL-9). Furthermore, both IL-1a and MIP-1b positively correlated with the non-ANA autoantibodies against MPO. Conversely, none of the 37 elevated inflammatory mediators were found to be associated with anti-PM/Scl-100 autoantibodies (data not shown).

We also analyzed the association of the inflammatory mediators with RBC/reticulocyte-leukocyte aggregation. Two chemokines (CCL23 and MIP-1a) and the growth factor HGF exhibited positive associations with the percentage of RBC/reticulocyte-leukocyte aggregates (Table 4). Together, our data demonstrate that inflammation is closely linked to autoimmunity and RBC aggregation with leukocytes in SCD participants at the steady state.

### Correlation analysis uncovered significant associations between inflammatory mediators and autoantibodies with VOCs/pain/sensory sensitivity

To investigate potential underlying factors contributing to VOCs, we examined the relationships of the elevated 37 inflammatory mediators, 4 autoantibodies, or RBC/reticulocyte-leukocyte aggregates with the intervals before and after VOC episodes (Table 5). After being adjusted for age, gender, and SCD genotypes, 6 inflammatory mediators (Extaxin-2, GRO-α, IL-2, IL-6, IL-17A, and LIF) showed positive correlations with the time intervals after VOCs. Conversely, the inflammatory cytokine IL-18 and granzyme A exhibited negative associations with the time interval before VOCs. These results suggest that specific inflammatory mediators, namely granzyme A and IL-18, might play a crucial role in the early detection or development of VOCs.

**Table 5:**
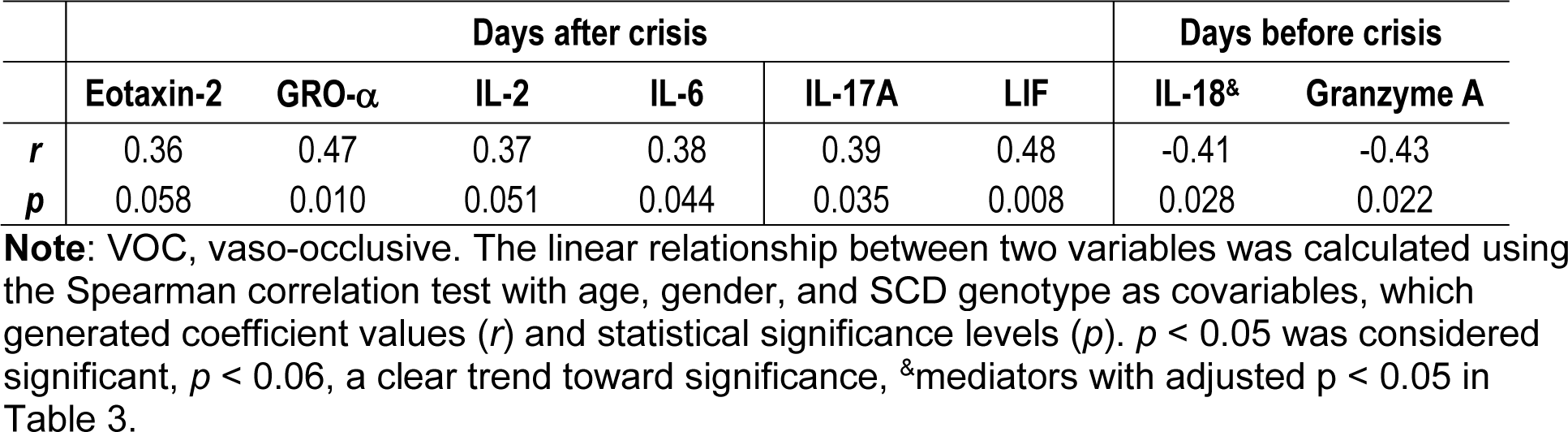
Correlations of altered inflammatory mediators with time interval from VOC crisis.

Next, we analyzed correlations of the inflammatory mediators with pain and sensory sensitivity as assessed by PROMs and QST. As presented in Table 6, several inflammatory mediators were associated with patient-reported pain intensity, physical function, and sensory sensitivity. A group of inflammatory mediators (G-CSF, granzyme A, BAFF, IL-9, and IL-17A) exhibited positive correlations with PedsQL scores. Conversely, most of the correlations between inflammatory mediators and various PROMs were negative. Specifically, 19 inflammatory mediators (CCL23, GM-CSF, GRO-a, MCP-2, Granzyme B, BAFF, bNGF, IL-7, IL-20, LIF, IL-2, IL-9, IL-17A, TNF-a, TNF-b, IL-5, IL-6, TSLP, and TREM-1) displayed negative associations with physical and psychological PROMs (Pain Intensity, Pain Episode, Depression Score, and Physical Function Score). Notably, 16 of the 19 factors were negatively correlated with the Physical Function Score, while 8 of them had negative correlations with Pain Episode Frequency/Recency. Moreover, 9 out of the 19 factors (Granzyme B, IL-20, LIF, IL-2, IL-17A, TNF-b, IL-5, IL-6, and TSLP) showed a positive association with the mechanical threshold (MDT forearm). Two additional factors, CCL21 and IL-8, were also positively associated with the MDT forearm. Five factors (Eotaxin-2, CCL21, GRO-a, MCP-2, and TREM-1) displayed positive correlations with pressure threshold (PPT trapezius). Furthermore, a group of inflammatory factors (Eotaxin-2, CCL21, GRO-a, MCP-2, MIP-3a, HGF, VEGF-A, IL-8, TNF-a, TNF-b, IL-5, and TREM-1) displayed negative correlations with mechanical pain threshold (MPT forearm and MTS forearm), and sensitivity threshold to cold (CPT forearm). Of note, several inflammatory factors, including GRO-a, MCP-2, granzyme B, IL-2, IL-17A, TNF-b, IL-5, and TREM-1, showed correlations with multiple measures from both PROMs and QST,

We also assessed the relationship of PROMs and experimental sensory sensitivity with the autoantibodies and RBC/reticulocyte-leukocyte aggregations (Table 6). We found that elevated autoantibodies against MPO exhibited a negative correlation with Physical Function Score and strong positive correlations with PPT (trapezius) and PPTol (trapezius). However, no correlations were detected between PROMS/QST and increased rate of RBC/reticulocyte-leukocyte aggregation, as well as BCAM and CD71 expression on the aggregates. Together, our results indicate that multiple inflammatory/autoimmune markers are inversely associated with the well-being of the SCD participants.

**Table 6.**
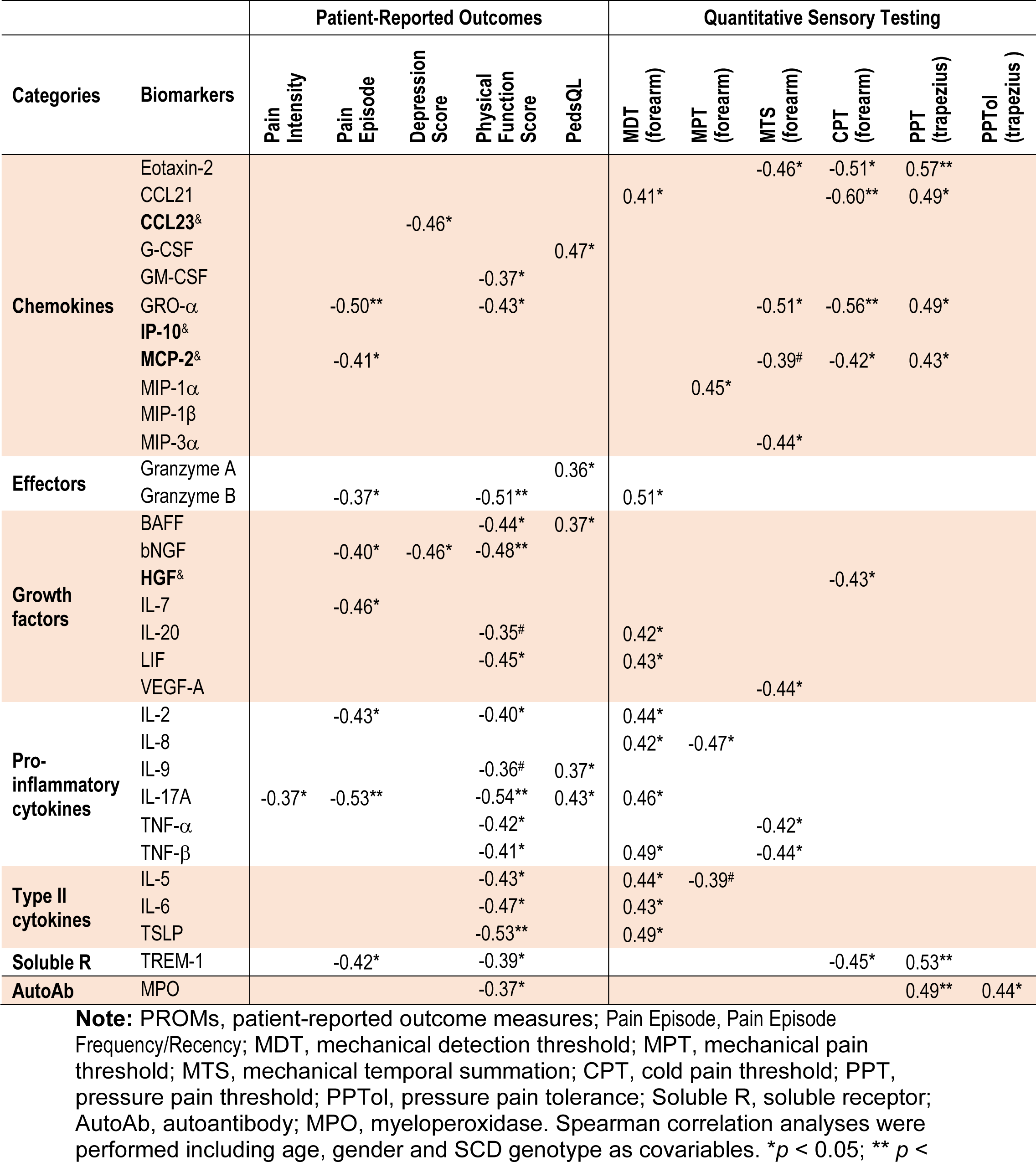
Correlations of upregulated soluble factors/autoantibody with PROMs/Sensory sensitivity in SCD subjects.

## Discussion

This study represents an interim analysis of an ongoing clinical trial that aims at investigating the clinical efficacy and neurobiological mechanisms of acupuncture analgesia in SCD participants (NCT05045820). Results showed that SCD participants in StSt showed higher levels of sensory sensitivity, as well as increased pain intensity, pain interference, widespread pain, physical dysfunction, and depression compared to HCs (Table 2). These results highlight the significant impact of SCD on patients’ physical and emotional well-being and align with previous studies on SCD (30, 34, 38, 39, 44, 45).

We collected cross-sectional blood samples and clinical parameters to comprehensively study inflammatory mediators, autoantibody profiles, and presence of RBC/reticulocyte-leukocyte aggregates, exploring their interrelations and potential relationship with VOCs, QoL, and sensory sensitivity. Specifically, we examined the plasma levels of 80 inflammatory mediators (Supplementary Table 1), the profiles of autoantibodies against 18 human antigens (Fig. 1), and the presence of RBC/reticulocyte-leukocyte aggregates in these participants (Fig. 2). Moreover, we investigated the correlations between these analytes and their potential relationship with VOCs, QoL, and sensory sensitivity in SCD participants. By examining these analytes and their relationships, we sought to identify potential mechanisms and therapeutic targets for further exploration and investigation.

SCD is characterized by a persistent pro-inflammatory state that leads to elevated levels of inflammatory mediators in the bloodstream. Inflammatory mediators are vital signaling molecules that play a crucial role in the development of pain, VOCs, and the pathogenesis of SCD (8, 17, 46). A recent study measured the serum levels of 27 inflammatory mediators including 15 cytokines, 7 chemokines, and 5 growth factors in 27 StSt and 53 HCs (28). The results revealed that significantly higher levels of several cytokines, chemokines, and growth factors in SCD participants compared to HCs (28). Strikingly, SCD participants exhibited elevated serum concentrations of pro-inflammatory molecules, such as IL-1β, IL-12p70, and IL-17A, when compared to SCD participants experiencing VOC (28). Here we expanded the spectrum of analyses and observed that SCD participants exhibited higher plasma levels of 37 out of 80 analytes, including 13 pro-inflammatory/anti-inflammatory cytokines, 10 chemokines, 10 growth factors, 2 effectors, and 2 soluble receptors (Table 3 and supplementary Tables 1 and 2). These findings collectively indicate a pro-inflammatory bias in SCD participants as compared to HCs.

Numerous cytokines elevated in our study, such as IL-1α, IL-4, IL-5,IL-6, IL-7, IL-8, TNF-a were previously shown to involved in VOC-related acute clinical complications such as acute chest syndrome, pulmonary hypertension, and pulmonary thrombosis (47-53). Systemic inflammation and modulation of the immune system with crosstalk of immunological molecules and immune cells were associated with the pathogenesis of persistent pain and the onset of VOCs in SCD. Plasma IFN-γ was augmented in steady-state SCD, which could modulate macrophage function and increase T helper cell expansion in SCD and most likely reflect inflammasome formation in inflammatory cells (52, 54, 55). Endothelium activation could in turn produce and release a number of potent inflammatory molecules, including IL-1α, IL-1β, IL-6, IL-8, GM-CSF, MCP-1, plasminogen activator inhibitor-1, and RANTES (52, 53, 56-58). The maturation and release of bioactive IL-1β and IL-18 (59-65), two vital immunoregulatory and proinflammatory cytokines, are governed by inflammatory caspases operating within specialized signaling platforms referred to as inflammasomes. Elevated plasma levels of IL-1β and IL-18 in SCD participants strongly suggest an aberration in the regulation of inflammasome activation within this particular group. In addition, IL-1β exerts a potent activation effect on leukocytes and ECs. Meanwhile, IL-18 plays an important role in stimulating vascular smooth muscle cell proliferation and migration, along with promoting the productions of IFN-γ, IL-2 and IL-12. These actions collectively exacerbate the inflammatory milieu in SCD participants (51, 54, 55). Notably, excessive production of IL-8 and RANTES contributes to the dehydration of sickle RBC, leading to an increase in RBC density and rigidity (66). This, in turn, enhances the adhesion of RBCs to the endothelium (67), a phenomenon closely linked to the severity of VOCs. These findings suggest that SCD participants experience persistently high levels of inflammatory mediators during StSt phases, even in the absence of severe clinical symptoms like VOCs.

Several inflammatory mediators were associated with the presence of autoantibodies and RBC/reticulocyte-leukocyte aggregation in SCD participants. Specifically, 8 inflammatory factors (MIP-1b, granzyme A, IL-34, VEGF-A, IL-1a, IL-9, IL-4, and IL-5) were positively associated with autoantibodies targeting 2 human antigens (SSA/Ro60 and MPO) (Table 4). Anti-SSA/Ro60 autoantibodies are typically classified as ANAs, while anti-MPO autoantibodies are categorized as non-ANAs. Thus, our results indicate that elevated inflammatory mediators are closely linked to both ANAs (anti-SSA/Ro60 autoantibodies) and non-ANAs (anti-MPO autoantibodies) in the bloodstream of SCD participants. Notably, the vast majority (7 out of 8) of inflammatory mediators displayed positive associations with both ANA (SSA/Ro60) and non-ANAs (Table 4). Given that mature RBCs in mammals lack nuclei, mitochondria, and other organelles (68), these inflammatory mediators are more likely to be implicated in autoimmune responses to cellular debris from other types of cells rather than mature RBCs. This finding suggests a potential link between ANA-associated autoimmunity and the presence of specific inflammatory mediators, which may contribute to our understanding of autoimmune processes and their association with certain cellular components in SCD.

In SCD participants, a complex and detrimental cycle is established through the interplay between inflammation and the formation of RBC/reticulocyte-leukocyte aggregates (69, 70). Inflammatory mediators disrupt immune responses, leading to an increased aggregation of RBCs/reticulocytes with leukocytes. Consequently, these aggregates exacerbate blood vessel blockages, further impeding blood flow and perpetuating the cycle of inflammation and tissue damage. This process triggers the release of additional inflammatory mediators, thus fueling a vicious circle of inflammation and VOCs. This vicious circle plays a significant role in the pathophysiology of SCD and significantly contributes to the recurrent pain crises and organ damage observed in affected individuals. We analyzed the association of inflammatory mediators with RBC/reticulocyte-leukocyte aggregation. In line with previous studies, our results also revealed 2 chemokines (CCL23 and MCP-1a) and 1 growth factor (HGF) that were associated with the aggregation (Table 4). These results provide important insights into the relationship between inflammation and RBC/reticulocyte-leukocyte aggregates that are associated with the occurrence and severity of VOCs.

SCD participants experience sudden and intense episodes of VOCs, which are challenging to predict and manage (71, 72). However, understanding of the circulating markers and their pathological processes during the transitioning phase from StSt to VOC episodes is extremely limited due to the unpredictable and rapid progression of the onset of VOCs. A previous study identified PDGF-BB and IL-1Rα as potential indicators for the acute-to-chronic stage in SCD (28). Our data revealed that 2 elevated inflammatory mediators (IL-18 and granzyme A) were associated with time the intervals prior to VOC onset and 6 mediators (Extaxin-2, GRO-α, IL-2, IL-6, IL-17A, and LIF) were associated with time intervals after active VOCs (Table 5), suggesting that these 8 mediators hold the potential to serve as novel biomarkers for predicting VOC episodes. Of particular note, IL-18 has emerged as a critical proinflammatory regulator in both innate and adaptive immune responses (73), and blocking IL-18 has been associated with mitigating neuropathic symptoms and enhancing the analgesic activity of morphine and buprenorphine (74). Inhibition of Granzyme A has been shown to reduce the levels of IL-6 and TNF-a (75). Thus, our results underscore the significance of specific inflammatory mediators in the early detection and development of VOCs, as well as potential avenues for pain management in SCD participants experiencing VOCs.

The existing literature has extensively studied the inflammation- and immunity-based mechanisms in SCD as compared to HCs, as well as the differences between StSt and VOC phases. However, there are limited studies investigating the relationship of inflammatory markers with clinical symptoms and sensory sensitivity during StSt. In the present study, we found that multiple inflammatory molecules were negatively associated with pain intensity (higher score indicated higher pain), the frequency/recency of clinical pain episodes (higher score indicated higher frequency/recency of acute pain episodes), emotional distress (depression), and physical dysfunction (higher score indicated higher physical dysfunction) in SCD participants. Conversely, higher levels of several soluble factors correlated with higher QoL and less pain. SCD participants with less severe Pain Episodes Score (higher score indicated higher severity) were associated with higher levels of GRO-a, MCP-2, granzyme B, bNGF, IL-7, IL-2, IL-17, and TREM 1 (Table 6). Of particular interest, higher IL-17A levels were also correlated with less pain intensity, physical dysfunction and higher QoL (higher PedsQL score indicated higher QoL). In fact, elevated IL-17A is associated with absence of acute chest syndrome in SCD participants, indicating a protective role (76). Consistently, higher levels of many inflammatory markers were also associated with decreased sensory sensitivity (in other words: increased threshold/tolerance to experimental stimuli) in response to mechanical (MDT, MPT, MTS), thermal (CPT), and pressure (PPT and PPTol) stimuli. We observed that SCD participants who had higher levels of Eotaxin-2, CCL21, GRO-a, MCP-2, HGF, and TREM-1 simultaneously exhibited lowered cold pain sensitivity (in other words: increased tolerance of cold-induced pain). These novel findings have not been reported in existing literature in SCD. The underlying mechanisms of these intriguing observations are unknown and warrant future investigation. We speculate that SCD participants experiencing recurrent extremely painful VOCs-related episodes developed resistance and pain inhibitory effect, thus exhibited decreased sensory sensitivity and increased tolerance to experimental pain stimuli, which therefore inhibited the antidromic release of proinflammatory mediators at periphery. This correlation profile suggested the interactive and distinct roles of inflammatory mediators in the processing of pain, and sensory sensitivity at StSt, and could be utilized for more extensive studies to explore the underlying nociceptive pathways in SCD.

In line with the correlation profile presented in Table 6, elevated anti-MPO autoantibodies were associated with decreased sensitivity and tolerance to pressure pain (PPT trapezius and PPTol trapezius). Notably, MPO plays a crucial role as a marker and modulator of inflammation and oxidative stress, primarily originating from activated leukocytes and neutrophils (77). A previous study demonstrated that increased MPO impairs EC function through vascular oxidative stress, while inhibiting MPO shows promise in improving vasodilation in mouse models of SCD (78). Furthermore, many SCD participants experience heightened cold sensitivity (79), and exposure to potential triggers, such as cold temperatures or stress, can induce vasoconstriction, potentially leading to VOC (80, 81). Therefore, elevated autoantibodies, such as anti-MPO autoantibodies (Fig.1), might contribute to vasoconstriction during the prodromal phase of VOCs while simultaneously interacting with immune- and inflammatory-targets (Table 4) to modulate pain inhibitory effect at StSt.

Inflammation, autoimmunity, and RBC/reticulocyte-leukocyte aggregations are believed to play a significant role in the pathogenesis of pain and VOCs in SCD participants (23, 82, 83). However, the direct connections of these factors with pain and VOCs in SCD participants are not fully understood. Our comprehensive analyses of these cross-sectional clinical samples provide valuable insights into the correlations between inflammatory mediators, autoantibody profiles, RBC/reticulocyte-leukocyte aggregation, clinical lab test results, and their associations with the onset of VOCs and pain sensitivity in SCD participants. This knowledge has the potential to significantly contribute to the development of suitable biomarkers/endpoints for clinical diagnosis of pain episodes in SCD participants, as well as targeted therapeutic approaches and improved management strategies for individuals affected by this condition. However, it is essential to acknowledge the limitations of our cross-sectional study, including small sample size, potential bias, confounding factors, the absence of insight into temporal trends, and the inability to establish causality. To address these limitations, we have undertaken efforts to collect longitudinal clinical samples throughout the duration of our clinical trial cohort and expand our sample size with ongoing recruitment efforts. This longitudinal approach is expected to provide a stronger foundation for establishing causal relationships between the analyzed variables. By observing changes over time, we can better infer cause-and-effect relationships of these variables, thus advancing our understanding of the mechanisms underlying the pathogenesis of SCD. Furthermore, the combination of cross-sectional and longitudinal approaches will enable us to gather complementary data, extending our investigations to address a broader range of clinical questions in both SCD participants and SCD participants experiencing acute VOC. The information gathered from these approaches will be pivotal in evaluating the clinical efficacy and neurobiological mechanisms of acupuncture analgesia in SCD participants participating in our clinical trial.

## Data Availability

All data produced in the present study are available upon reasonable request to the authors

https://medicine.iu.edu/faculty/51903/wang-ying

## Author contribution

W.L., Q.Y., and Y.W. supervised experimental performance and data collection, analysis, and interpretation; A.Q.P., C.D., and B.R. assisted with data collection and figure preparation. F.S. assisted with data analyses and figure preparation; W.L., Q.Y., and Y.W. drafted and edited the manuscript; S.E.H., and R.E.H. helped with editing the manuscript; Y.W. and A.R.O. directed patient recruitment; A.R.O., R.M., N.M., S.A.J., B.M.H., A.G. facilitated recruitment for the study; Y.W. developed the concepts, directed overall performance and quality of the clinical investigation.

## Authors Statement of Competing Interest

All authors have read the journal’s authorship agreement and policy on disclosure of potential conflicts of interest.

## Acknowledgement

The authors would like to thank Tyler James Barret, Nayana Dutt, Payton Mittman, Bea Paras, Ramat Gbemisola Suleiman-Oba, Amy Gao and Yongqi Yu for assisting with experimental performance, and clinical team staff at Indiana University Clinical Research Center for patient scheduling and blood draw performance.

## Funding Support

This work was supported by NIH K99/R00 award (Grant # 4R00AT010012 to Y.W.) and Indiana University Health – Indiana University School of Medicine Strategic Research Initiative funding to Y.W.

## Abbreviations

α4β1: alpha-4 beta-1
ANA: anti-nuclear autoantibody
CDT: cold detection threshold
CPT: cold pain threshold
CPM: conditioned pain modulation
CRP: C-reactive protein
DC: dendritic cell
EC: endothelial cell
GPA: glycophorin A
HADS: Hospital Anxiety and Depression Scale
HC: healthy control
HCT: hematocrit
HDT: heat detection threshold
Hgb: hemoglobin
HPT: heat pain threshold
LIF: leukemia inhibitory factor
Lu/BCAM: lutheran/basal cell adhesion molecule
MCV: mean corpuscular volume
MCH: mean corpuscular hemoglobin
MDT: mechanical detection threshold
MFI: median fluorescent intensity
MPO: myeloperoxidase
MPT: mechanical pain threshold
MTS: mechanical temporal summation
PBMC: peripheral blood mononuclear cell
PedsQL: Pediatric Quality of Life Inventory
PMN: polymorphonuclear neutrophil
PPT: pressure pain threshold
PPTol: pressure pain tolerance
PROMIS: Patient-Reported Outcomes Measurement Information System
PROMs: patient-reported outcome measures
QoL: quality of life
QST: quantitative sensory testing
RBC: red blood cell
RDW: red cell distribution width
SCD: sickle cell disease
SLE: systemic lupus erythematosus
SSA/Ro60: Sjögren’s Syndrome-related antigen A/Ro60 kDa
StSt: steady-state condition
TSLP: thymic stromal lymphopoietin
TSP: temporal summation of pain
VOC: vaso-occlusive crisis
WBC: white blood cell.

## Notes

**Conflict of interest** The authors declare no conflicts of interest that pertain to this manuscript.

### Competing Interest Statement

The authors have declared no competing interest.

### Clinical Trial

NCT05045820

### Clinical Protocols

https://classic.clinicaltrials.gov/ct2/show/NCT05045820

### Funding Statement

NIH (K99/R00 Grant # 4R00AT010012 to Y.W.)

### Author Declarations

This study was performed with the approval of the Institutional Review Boards (IRB) at Indiana University School of Medicine, and each participant provided written informed consent during the screening visit prior to the subsequent study procedure.

